# The potential of low molecular weight heparin to mitigate cytokine storm in severe COVID-19 patients: a retrospective clinical study

**DOI:** 10.1101/2020.03.28.20046144

**Authors:** Chen Shi, Cong Wang, Hanxiang Wang, Chao Yang, Fei Cai, Fang Zeng, Fang Cheng, Yihui Liu, Taotao Zhou, Bin Deng, Israel Vlodavsky, Jin-Ping Li, Yu Zhang

## Abstract

**Background:** On March 11, 2020, the World Health Organization declared its assessment of COVID-19 as a global pandemic. However, specific antiviral drugs are still unavailable, and pateints are managed by multiple complementary treatments.

**Methods:** The electronic medical records of COVID-19 patients where basic information, complete blood count, coagulation profile, inflammatory cytokines and serum biochemical indicators in 42 patients with COVID-19 (21 of whom were treated with low molecular weight heparin (LMWH), and 21 without LMWH) that were retrospectively analyzed to compare and evaluate the effect of LMWH treatment on disease progression.

**Findings:** 42 patients with COVID-19 treated at the hospital between February 1 and March 15, 2020, were selected for the study, of which 21 underwent LMWH treatment (LMWH group), and 21 did not (Control), during hospitalization. Changes in the percentage of lymphocytes in the LMWH group before and after LMWH treatment were significantly different from those in the control group (11·10±9·50 vs. 3·08±9·66, *p*=0·011, respectively). Changes in the levels of D-dimer and fibrinogen degradation products (FDP) in the LMWH group before and after LMWH treatment were significantly different from those in the control group (-2·85±3·90, -0·05±0.85, *p*=0·002; -9·05±13·14, -1·78±3·15, *p*=0·035). Strikingly, in the LMWH group, IL-6 levels were significantly reduced after LMWH treatment (47·47±58·86, 15·76±25·71, *p*=0·006). Besides, the changes in IL-6 levels in the LMWH group before and after LMWH treatment were significantly different from those in the control group (-32·46±65·97, 14·96±151·09, *p*=0·031).

**Interpretation:** LMWH improves the coagulation dysfunction of COVID-19 patients and exerts anti-inflammatory effects by reducing IL-6 and increasing lymphocyte %. It appears that LMWH can be used as a potential therapeutic drug for the treatment of COVID-19, paving the way for a subsequent well-controlled clinical trial.

**Funding:** National Natural Science Foundation of China (No. 81603037 to SC) and the National Key Research and Development Plan of China(2017YFC0909900).

## Introduction

On March 11, 2020, the World Health Organization (WHO) declared its assessment of COVID-19 as a global pandemic. SARS-CoV-2 is characterized by a long incubation period, high infectivity, and multiple routes of transmission.^1,2^ According to real-time WHO statistics, the total number of confirmed cases of COVID-19 worldwide as of April 6, 2020 has exceeded a million with more than 70000 deaths. However, no effective medicines are currently available, so patients are treated symptomatically. Given the rapid spread of COVID-19 and the high mortality rate in severe cases, there is an urgent need to promptly control the occurence of a severe disease. A better understanding of the mechanisms of pathological changes will help to screen reliable drugs out of presently existing medications.

Lymphopenia and inflammatory cytokine storm are typical abnormalities observed in highly pathogenic coronavirus infections (such as SARS and MERS),^3^ believed to be associated with disease severity.^4-6^ Several clinical studies revealed that cytokine storms are important mechanisms underlying disease exacerbation and death of COVID-19 patients.^4-6^ Particularly, IL-6 levels in severely ill patients were significantly higher than in mild cases.^7^ IL-6 is one of the core cytokines that are consistently found to be elevated in the plasma of patients with cytokine storm,^8^ contributing to many of the key symptoms of cytokine storm, such as vascular leakage, activation of the complement and coagulation cascades, inducing disseminated intravascular coagulation (DIC).^9,10^ Reducing the level and activity of IL-6 may contribute to prevent or even reverse the cytokine storm syndrome,^11^ thereby improving the condition of patients with COVID-19.

Substantial studies have reported that low molecular weight heparin (LMWH) has various non-anticoagulant properties^12^ that play an anti-inflammatory role by reducing the release of IL-6.^13-15^ However, the anti-inflammatory effects of LMWH in COVID-19 are currently unknown. By analyzing the relieving effect of LMWH in patients with COVID-19 our retrospective cohort study demonstrates, for the first time, the significant benifiical effect of LMWH in controlling ctytokine storm. This approach is believed to delay disease progression in COVID-19 patients, strongly encuraging a well-controlled clinical practice (**Fig. 1**).

**Figure 1.**
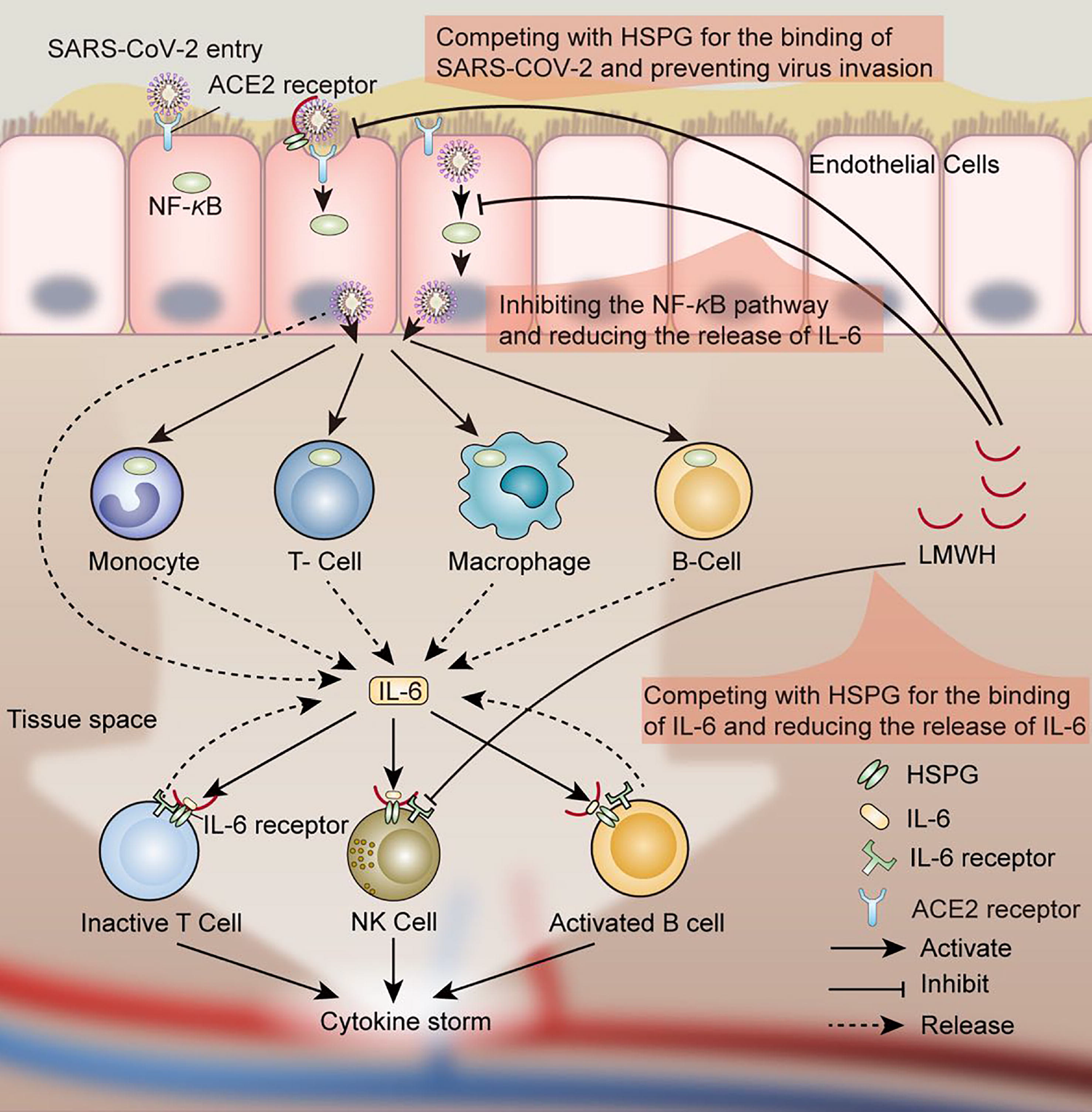
Possible mechanism of anti-inflammatory effects of LMWH in patients with COVID-19. Under conventional antiviral treatment regimens, LMWH improves hypercoagulability, inhibits IL-6 release, and attenuates IL-6 biological activity. It has potential antiviral effects and helps delay or block inflammatory cytokine storms. LMWH can increases the LYM% in the patients. The multiple effects of LMWH encourages its application for the treatment of COVID-19 patients.

## Methods

### Research subjects

To investigate the therapeutic effect of LMWH on COVID-19, we conducted a retrospective study. All cases in this study were located at Union Hospital, Tongji Medical College, Huazhong University of Science and Technology (Wuhan, Hubei Province, China), a designated treatment hospital for patients with COVID-19. This study was approved by the institutional review board of the hospital. In total, we retrospectively collected the electronic medical records of 42 patients with COVID-19, the admission data for these patients was from February 1, 2020, to March 15, 2020 (**Fig. 2** shows the case inclusion flowchart), of which 21 underwent LMWH treatment (defined as LMWH group), and 21 did not (defined as Control group) (**Table 1**), during hospitalization.

**Table 1.** LMWH use in treating conditions of the 21 patients with COVID-19. Details of the dose, frequency, route of administration, and days of use of LMWH in the LMWH group.

| LMWH Group<br>(n=21) | Treatment with LMWH | Days of treatment |
| --- | --- | --- |
| H1 | Enoxaparin sodium injection 4000AxaIU qd i·h· | 10 |
| H2 | Enoxaparin sodium injection 4000AxaIU qd i·h· | 10 |
| H3 | Enoxaparin sodium injection 4000AxaIU qd i·h· | 14 |
| H4 | Enoxaparin sodium injection 4000AxaIU qd i·h· | 13 |
| H5 | Enoxaparin sodium injection 4000AxaIU qd i·h· | 17 |
| H6 | Nadroparin calcium injection 4100AxaIU qd i·h· | 9 |
| H7 | Enoxaparin sodium injection 4000AxaIU qd i·h· | 2 |
| H8 | LMWH sodium injection 5000IU once i·h· | 1 |
| H9 | Enoxaparin sodium injection 4000AxaIU qd i·h· | 16 |
| H10 | Enoxaparin sodium injection 4000AxaIU qd i·h· | 19 |
| H11 | Enoxaparin sodium injection 4000AxaIU qd i·h· | 14 |
| H12 | Enoxaparin sodium injection 2000AxaIU qd i·h· | 19 |
| H13 | Enoxaparin sodium injection 2000AxaIU qd i·h· | 22 |
| H14 | Nadroparin calcium injection 4100AxaIU qd i·h· | 11 |
| H15 | Nadroparin calcium injection 4100AxaIU qd i·h· | 13 |
| H16 | Enoxaparin sodium injection 4000AxaIU qd i·h· | 8 |
| H17 | Nadroparin calcium injection 4100AxaIU qd i·h· | 19 |
| H18 | LMWH sodium injection 5000IU qd i·h· | 8 |
| H19 | Enoxaparin sodium injection 4000AxaIU qd i·h· | 8 |
| H20 | Enoxaparin sodium injection 4000AxaIU qd i·h· | 7 |
| H21 | Enoxaparin sodium injection 4000AxaIU qd i·h· | 10 |

**Figure 2.**
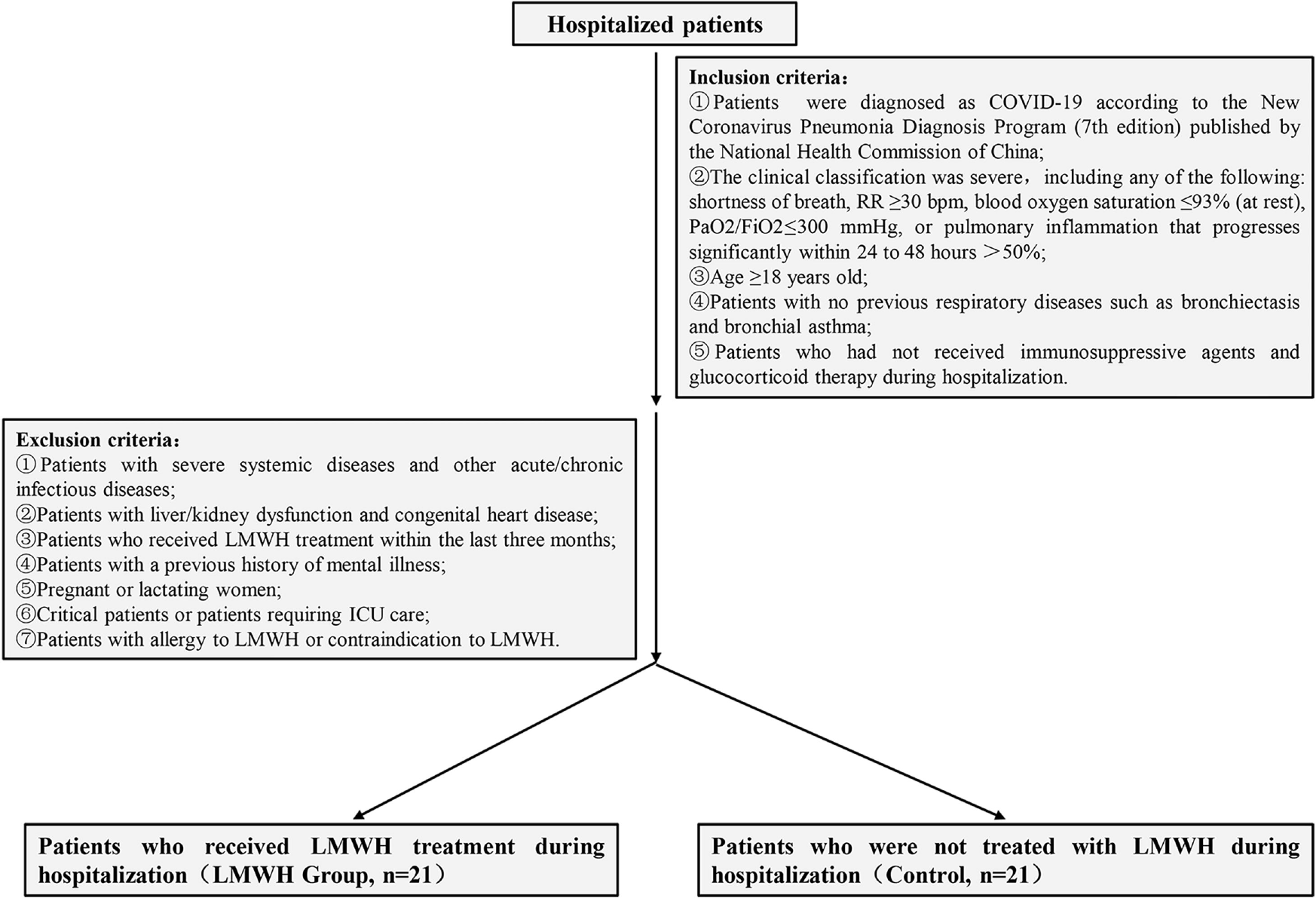
Flowchart of inclusion and exclusion criteria for patients with COVID-19. Based on strict inclusion and exclusion criteria, 42 patients with COVID-19 treated at the hospital between February 1 and March 15, 2020, were selected for the study, of which 21 underwent LMWH treatment (LMWH group) and 21 did not (Control group) during hospitalization.

Inclusion criteria: (1) met the diagnostic standards of novel coronavirus pneumonia (7th edition) formulated by the National Health Commission of China; (2) experienced any of the following: shortness of breath, respiration rate(RR) ≥ 30 breaths/minute; resting oxygen saturation ≤ 93%; PaO2/FiO2 ≤ 300 mmHg; lung imaging showing significant lesion progression of > 50% within 24-48 h, and a severe clinical classification; (3) age ≥ 18 years; (4) no previous history of bronchiectasis, bronchial asthma, or other respiratory diseases; (5) no immunosuppressant or glucocorticoid use during treatment.

Exclusion criteria: (1) patients with severe systemic diseases and other acute or chronic infectious diseases; (2) patients with liver and kidney insufficiency or congenital heart disease; (3) patients who had been treated with LMWH in the previous three months; (4) patients with a prior history of mental illness; (5) pregnant or lactating women; (6) patients clinically classified as critically ill or housed in the intensive care unit (ICU); (7) patients allergic to LMWH or contraindicated for LMWH.

### Data collection

The basic information, complete blood count, coagulation profile, inflammatory cytokines, and serum biochemical indicators (including liver function, kidney function, lactate dehydrogenase, C-reactive protein (CRP) and electrolytes) of 42 patients with COVID-19 were retrospectively analyzed. Two researchers also independently reviewed the data collection forms to double-check the data collected.

### Statistical analysis

Data analysis was performed using SPSS 22·0 statistical software. Data are expressed as mean ± standard deviation (SD). GraphPad 6·0 software was used for plotting. Differences between groups were evaluated using the T-test for measurement data, the Chi-square test for count data, and the Kruskal-Wallis nonparametric test between groups (independent samples) and within groups (related samples). Differences of *p* < 0·05 were considered statistically significant.

## Results

### General characteristics of patients with COVID-19

As shown in **Table 2**, the LMWH group consisted of 13 males and eight females aged between 42 and 91 years (median age = 69·0 years), and the Control group consisted of 14 males and seven females aged between 40 and 84 years (median age = 69·0 years). There were no significant differences in comorbidities, such as hypertension, diabetes, cardiovascular disease, and cancer, between the two groups. Similarly, there were no significant differences in coronavirus pneumonia onset symptoms, including fever (body temperature ≥ 37·3°C), cough, sputum, chest distress or asthma, myalgia, fatigue, anorexia, diarrhea, and nausea and vomiting. Similarly, there was no significant difference in antiviral treatment between the two groups. These results indicate that the general characteristics of the two groups of patients were consistent and comparable.

**Table 2.** General characteristics of all the enrolled patients with COVID-19. There were no significant differences in age, sex, comorbidities, onset symptoms, time from hospitalization to virus shedding, length of hospital stay, antiviral treatment, and disease progression between the two groups. Data are median (IQR) or n(%). p values are for comparing the LMWH group and Control group. NA = not applicable.

|  | LMWH Group<br>(n=21) | Control<br>(n=21) | p value |
| --- | --- | --- | --- |
| Characteristics |  |  |  |
| Age, years | 69·0(42·0-91·0) | 69·0(40·0-84·0) | 0·54 |
| Sex | - | - | 0·75 |
| Female | 8(38%) | 7(33%) | - |
| Male | 13(62%) | 14(67%) | - |
| Comorbidity | 13(62%) | 8(38%) | 0·12 |
| Hypertension | 8(38%) | 5(24%) | 0·32 |
| Diabetes | 6(29%) | 2(10%) | 0·12 |
| Cardiovascular disease | 5(24%) | 2(10%) | 0·21 |
| Chronic obstructive lung disease | 0 | 0 | NA |
| Carcinoma | 0 | 1(5%) | 0·31 |
| Chronic kidney disease | 0 | 0 | NA |
| Other | 4(19%) | 1(5%) | 0·15 |
| Signs and symptoms |  |  |  |
| Fever (temperature $\geq 37.3^{\circ}\text{C}$) | 15(71%) | 13(62%) | 0·51 |
| Cough | 9(43%) | 7(33%) | 0·53 |
| Sputum | 6(29%) | 4(19%) | 0·47 |
| Chest distress or asthma | 11(52%) | 8(38%) | 0·35 |
| Myalgia | 2(10%) | 3(14%) | 0·63 |
| Fatigue | 8(38%) | 5(24%) | 0·32 |
| Anorexia | 6(29%) | 5(24%) | 0·73 |
| Diarrhoea | 2(10%) | 1(5%) | 0·55 |
| Nausea or vomiting | 2(10%) | 1(5%) | 0·55 |
| Respiratory rate $\geq 30$ breaths per min | 0 | 0 | NA |
| Pulse $\geq 125$ beats per min | 0 | 0 | NA |
| Systolic blood pressure $< 90$ mmHg | 0 | 0 | NA |
| Antiviral therapy |  |  |  |
| Arbidol | 18(86%) | 20(95%) | 0·29 |
| Recombinant Human Interferon $\alpha 2\text{B}$<br>(aerosol inhalation) | 6(29%) | 6(29%) | 1·00 |
| Ribavirin | 2(10%) | 0 | 0·15 |
| Lopinavir/Ritonavir | 2(10%) | 0 | 0·15 |
| Traditional Chinese medicine decoction | 11(52%) | 9(43%) | 0·54 |
| Disease progression |  |  |  |
| Improved | 21(100%) | 21(100%) | NA |
| Invariable | 0 | 0 | NA |
| Deteriorative | 0 | 0 | NA |
| Time from hospitalization to virus shedding<br>after the onset of the COVID-19, days | 20·0(11·0-31·0) | 19·0(12·0-30·0) | 0·46 |
| Hospital length of stay, days | 29·0(17·0-42·0) | 27·0(24·0-31·0) | 0·41 |

### LMWH has no effect on the duration of conversion to negative and the length of patient hospitalization

As shown in **Table 2**, the number of days to convert virus to negative (time from admission to virus shedding) was 20·0 days (IQR 11·0-31·0) in the LMWH group and 19·0 days (IQR 12·0-30·0) in the Control group (*p* = 0·46); the difference between the two groups was not significant. Similarly, the length of hospital stay was 29·0 days (IQR 17·0-42·0) in the LMWH group and 27·0 days (IQR 24·0-31·0) in the Control group (*p* = 0·41); the difference between the two groups was not significant. Notably, all patients in the LMWH group and the Control group showed overall improvement after treatment.

### Effect of LMWH on blood routine characteristics

As shown in **Fig.3**A-D, there was no significant difference in red blood cells (RBC), white blood cells (WBC), monocyte%, and neutrophil% levels between the two groups. **Fig. 3**E reveals no significant difference in lymphocyte% between the LMWH and Control groups before LMWH treatment (18·84±8·24, 22·42±8·74, *p*=0·144). There was also no significant difference in lymphocyte% between the LMWH and Control groups after LMWH treatment (29·94±7·92, 25·65±10·10, *p*=0·215). However, patients in the LMWH group had a significantly increased percentage of lymphocytes after LMWH treatment (18·84±8·24, 29·94±7·92, *p*=0·00048). Besides, the changes in lymphocyte% in patients of the LMWH group before and after LMWH treatment were significantly different from those in the Control group (11·10±9·50, 3·08±9·66, *p*=0·011).

**Figure 3.**
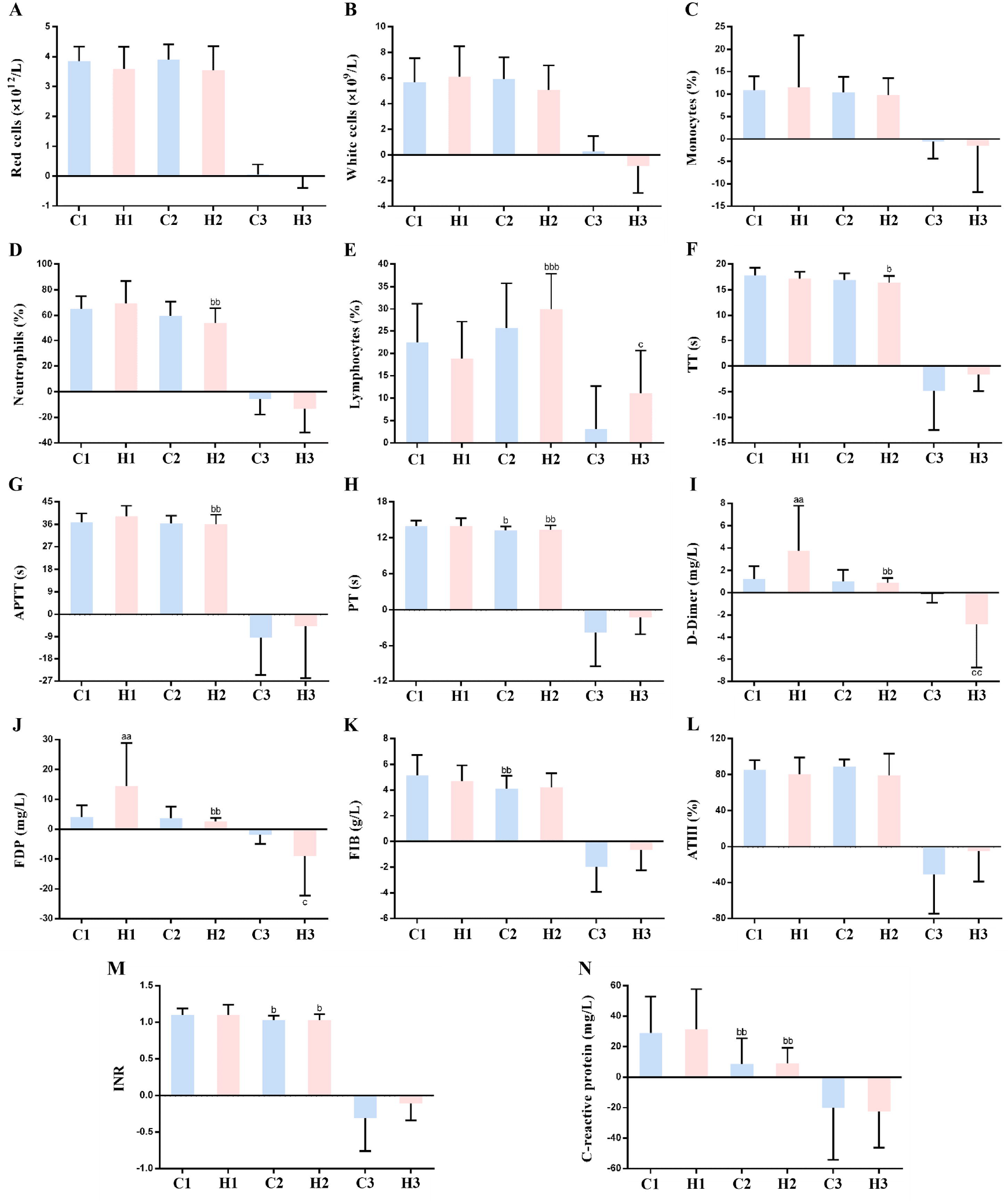
Effect of LMWH on complete blood count, coagulation profile, and CRP in the enrolled patients with COVID-19. **A-N:** Red blood cells (A), white blood cells (B), monocytes% (C), neutrophils% (D), lymphocytes% (E), TT (F), APTT (G), PT (H), D-dimer (I), FDP (J), FIB (K), AT III (L), INR (M) and CRP (N) levels in patients with COVID-19. Data are expressed as mean ± standard deviation (SD) (n = 21). C1 vs. H1 or C2 vs. H2, ^a^ *p* < 0·05, ^aa^ *p* < 0·01, ^aaa^ *p* < 0·001; C1 vs. C2 or H1 vs. H2, ^b^ *p* < 0·05, ^bb^ *p* < 0·01, ^bbb^ *p* < 0·001; C3 vs. H3, ^c^ *p* < 0·05, ^cc^ *p* < 0·01, ^ccc^ *p* < 0·001. (C1: Control group, indices at admission; C2: Control group, indices at discharge; C3: Control group, changes in indices during hospitalization; H1: LMWH group, indices before LMWH treatment; H2: LMWH group, indices after LMWH treatment; H3: LMWH group, changes in indices before and after LMWH treatment.).

### Effect of LMWH on coagulation parameters

There was no significant difference in thrombin time (TT, **Fig. 3**F), activated partial thromboplastin time (APTT, **Fig. 3**G), and prothrombin time (PT, **Fig. 3**H) levels between the two groups. As shown in **Fig. 3**I, the levels of D-dimer in the LMWH group were significantly higher compared to those in the Control group before treatment (3·75±4·04, 1·23±1·15, *p*=0·009). There was no significant difference in D-dimer levels between the LMWH and Control groups after LMWH treatment (0·90±0·44, 1·00±1·06, *p*=0·368). Upon LMWH treatment, the D-dimer levels were significantly reduced in the LMWH group (3·75±4·04, 0·90±0·44, *p*=0·001) (**Fig. 3**I). The changes in D-dimer levels in patients in the LMWH group before and after LMWH treatment were significantly different from those in the Control group (-2·85±3·90, -0·05±0·85, *p*=0·002). As shown in **Fig. 3**J, the levels of fibrinogen degradation products (FDP) in the LMWH group were significantly higher compared to those in the Control group before treatment (14·35±14·6, 4·05±3·9, *p*=0·002). There was no significant difference in FDP levels between the LMWH and Control groups after LMWH treatment (2·64±1·16, 3·59±4·00, *p*=0·959). In the LMWH group, FDP levels were significantly reduced after LMWH treatment (14·35±14·6, 2·64±1·16, *p*=0·001). The changes in FDP levels in patients of the LMWH group before and after LMWH treatment were significantly different from those in the Control group (-9·05±13·14, -1·78±3·15, *p*=0·035). However, there was no significant difference in fibrinogen (FIB, **Fig. 3**K), antithrombin (ATIII, **Fig. 3**L) and international normalized ratio (INR, **Fig. 3**M) levels between the two groups.

### Effect of LMWH on CRP levels

As shown in **Fig. 3**N, LMWH treatment had no significant effect on CRP levels. There is no difference between the two groups of patients before LMWH treatment (31·15±26·62, 29·00±23·79, *p*=0·497), nor after LMWH treatment (8·95±10·44, 8·76±16·66, *p*=0·620). Consequently, there were no significant differences in the changes in CRP levels between the two groups of patients before and after LMWH treatment (-22·62±23·79, -20·23±33·91, *p*=0·660).

### Effect of LMWH on cytokine levels in patients with COVID-19

Finally, we have analyzed the levels of inflammatory cytokines in the two groups. As shown in **Fig. 4**, there were no significant differences in the levels of IL-2, IL-4, IL-10, TNF-α, and IFN-γ between the LMWH treated and untreated groups. Notably, application of LMWH significanlty lowered the level of IL-6. As shown in **Fig. 4**B, both groups had a high level of IL-6, and there was no significant difference between the LMWH and Control groups before treatment (47·47±58·86, 63·27±96·27, *p*=0·950). In contrast, after LMWH treatment, the levels of IL-6 in the LMWH group were significantly lower compared to those in the Control group (15·76±25·71, 78·24±142·41, *p*=0·00039). Accordingly, IL-6 levels in the LMWH group were significantly reduced after LMWH treatment (47·47±58·86, 15·76±25·71, *p*=0·006). Similarly, the changes in IL-6 levels in the LMWH group before and after LMWH treatment were significantly different from those in the control group (-32·46±65·97, 14·96±151·09, *p*=0·031).

**Figure 4.**
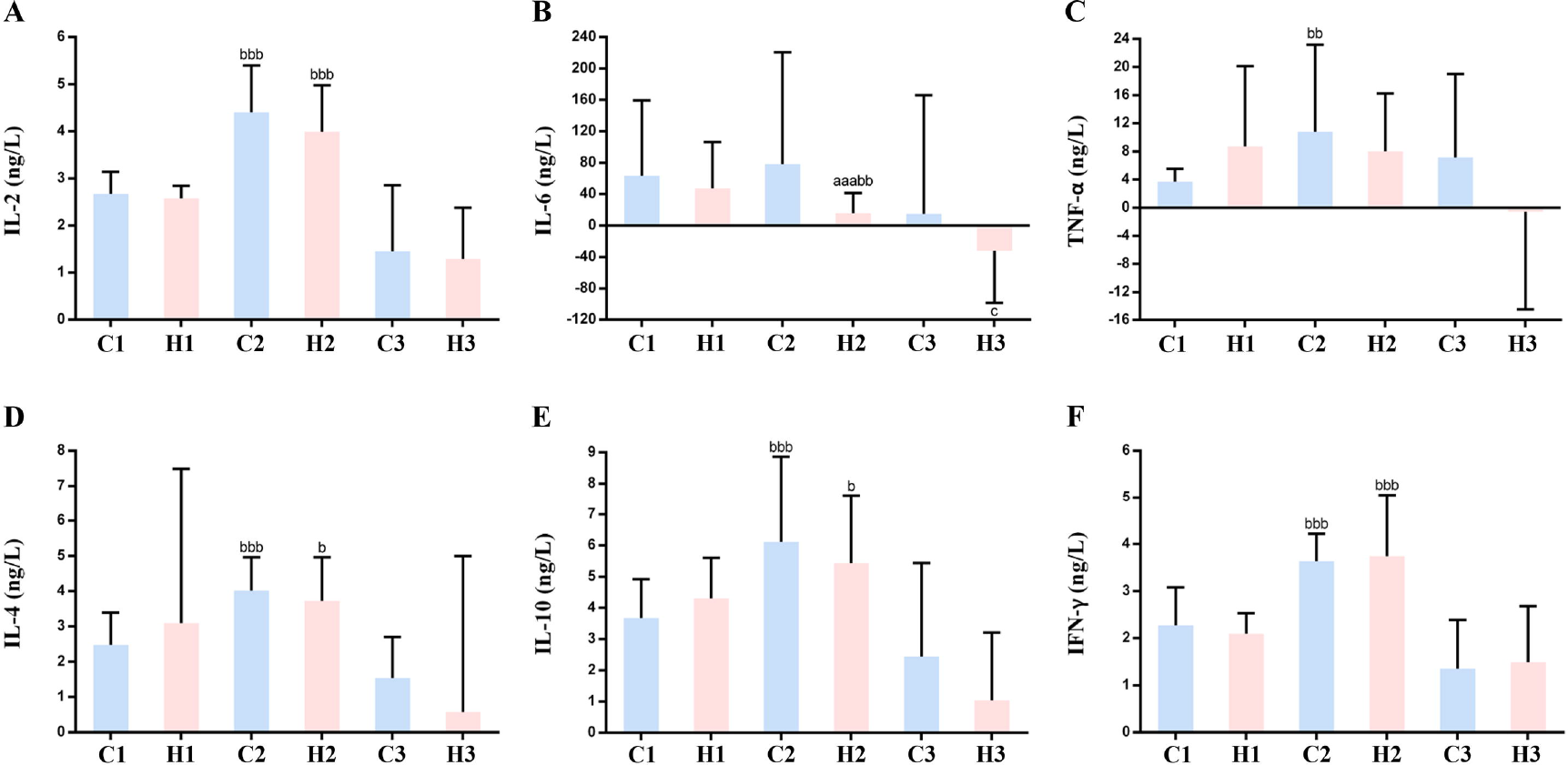
Effect of LMWH on inflammatory cytokines in the enrolled patients with COVID-19. **A-F:** IL-2 (A), IL-6 (B), TNF-α (C), IL-4 (D), IL-10 (E), and IFN-γ (F) levels in the two groups of patients with COVID-19. Data are expressed as mean ± standard deviation (SD) (n = 21). C1 vs. H1 or C2 vs. H2, ^a^ *p* < 0·05, ^aa^ *p* < 0·01, ^aaa^ *p* < 0·001; C1 vs. C2 or H1 vs. H2, ^b^ *p* < 0·05, ^bb^ *p* < 0·01, ^bbb^ *p* < 0·001; C3 vs. H3, ^c^ *p* < 0·05, ^cc^ *p* < 0·01, ^ccc^ *p* < 0·001. (C1: Control group, indices at admission; C2: Control group, indices at discharge; C3: Control group, changes in indices during hospitalization; H1: LMWH group, indices before LMWH treatment; H2: LMWH group, indices after LMWH treatment; H3: LMWH group, changes in indices before and after LMWH treatment).

## Discussion

Cytokine storms are associated with deterioration in several infectious diseases, including SARS and avian influenza,^3,16^ and are an important cause for exacerbation in patients.^17^ In recent years, studies have revealed that heparin has various non-anticoagulant properties, for example, LMWH can exert anti-inflammatory effects by reducing the release of IL-6.^13-15,18^

It was reported that IL-6 and IL-8 could cause hypercoagulation, leading to scattered fibrin clots, shortening the clot dissolution time and maximizing the dissolution rate.^19^ It was also observed that severe COVID-19 patients had higher levels of IL-6,^7^ suggesting that the hypercoagulation status of COVID-19 patients may be related to the elevated levels of cytokines. In previous studies of patients with COVID-19, D-dimer levels were significantly elevated in patients admitted to the ICU with severe disease.^20^ Higher levels of D-dimer and FDP in fatal cases have been reported,^21^ and there was certain correlation between D-dimer and COVID-19 severity.^22^ However, there is currently no conclusive evidence supporting the use of D-dimer as an evaluation index.^23-25^ A broad sample analysis is required to determine whether D-dimer is associated with COVID-19 severity. Therefore, the present study does not consider this parameter as an evaluation index for disease progression. The average values of D-dimer and FDP before treatment was higher in the LMWH group than in the control group (3·75, 1·23, *p*< 0·01; 14·35, 4·05, *p*<0·01), therefore LMWH was applied. Because this trial is a retrospective analysis, we did not intervene in the type of treatment given to the patients, inferring that the purpose of medication in the LMWH group was to improve hypercoagulability. Because D-dimer and FDP are not considered as factors that designate patient’s disease progression, their levels had no effect on subsequent analysis of the results.

Apart of its anticoagulant activity, there are other routes to explain a favorable effect of LMWH on COVID-19 patients. Heparan sulfate (HS), a linear polyanionic polysaccharide, is a major constituent of all mammalian cells and tissues.^26^ It highly resembles heparin and LMWH in its structural properties and sugar composition.^26^ Importantly, HS has been known to serve as the first point of contact between target cells and a large number of human viruses (i.e., dengue virus, hepatitis C virus, human immunodeficiency virus, human papilloma virus, herpes viruses),^27,28^ including the SARS-CoV-2 virus.^29,30^ A very recent online paper has used surface plasmon resonance and circular dichroism and showed that the SARS-CoV-2 Spike S1 protein receptor binding domain interacts with heparin.^30^ Heparin, LMWH and heparin-like compounds have been shown to efficiently compete with HS and thereby attenuate viral attachment and infection,^31^ providing a straightforward explanation for the anti-viral effect of LMWH in clinical settings.

Importantly, as clarified below, recent studies expanded the established role of HS from a viral attachment molecule to an essential receptor required for entry. Heparin, LMWH and non-anticoagulants species of heparin are known to inhibit the enzymatic activity of heparanase,^32^ the sole HS-degrading endoglycosidase, shown recently to promote viral infection and spread.^33-35^ It appears that heparanase behaves as a molecular switch in viral infection, which transforms the cell from a virus-permissive mode in which viral attachment and entry are favored, to a virus-deterring mode which allows for viral detachment and egress from cells.^33^ Briefly, it was found that upregulation and activation of heparanase is a strategy common to a broad range of viral species (i.e., PRRSV, vaccinia virus) to increase egress, spread and transmission.^35^ Interestingly, it appears that heparanase plays a role also in driving the undesirable cytokine storm discussed above. In individuals with SARS-CoV-2 infection, the level of inflammatory cytokines is markedly higher than normal and held responsible for the severity of the disease. Agelidis et al. documented that upon HSV-1 infection, heparanase translocate to the nucleus of the infected cells and promotes inflammatory signaling, mediated primarily via NF-κB.^35^ In fact, transcription of IL-6 was significantly decreased after treatment with an inhibitor of heparanase enzymatic activity.^35^ LMWH which inhibits heparanase activity^32^ may have a similar effect, possibly providing a mechanistic explanation for the decrease in IL-6 that we observed in the LMWH treated patients. Collectively, the above considerations suggest that heparanase inhibitors (i.e., LMWH) may be an effective strategy in a therapeutic or combination therapy against viral infection, including COVID-19. Additional studies showed that inhibition of the glycocalyx-degrading enzymes sialidase, cathepsin L and heparanase, using a combination therapy of zanamavir, cathepsin-L and heparanase inhibitors, decreased vascular leakage after exposure to the influenza virus NS1 protein in vitro and in vivo.^36^ It will be interesting to see if an analogous therapeutic inhibition of glycocalyx breakdown can provide similar benefit clinically.

Several studies have recommended CRP and lymphocyte% (LYM%) as indices for evaluating the effectiveness of clinical drugs or treatments.^5,37,38^ In the various analyses applied in this study, there was no statistically significant difference in CRP levels between the groups, indicating that LMWH treatment has no effect on this parameter. Notably, LYM% was higher in the LMWH group compared to the Control group (*p* < 0·001), consistent with the results of Derhaschnig *et al*.^39^ This suggests that LMWH can increase LYM% in patients with COVID-19 and thereby improve their condition. Furthermore, it was reported that proinflammatory cytokines, such as TNFα and IL-6, can induce lymphopenia.^6^ Hence, the decrease in IL-6 (**Fig**. 4B) may contribute to the increase in LYM% observed in the LMWH treated patients.

IL-6 levels in severely ill patients with COVID-19 are significantly higher than in patients with mild disease.^7^ Transition from mild to severe conditions in patients with COVID-19 occur when cytokine levels reach and/or exceed a certain threshold, leading to a cytokine release syndrome.^8^ Hence, reducing IL-6 release is expected to attenuate the cytokine storm syndrome caused by the virus,^9^ thereby improving the condition of patients with COVID-19. LMWH was reported to reduce the release of IL-6 in the body by inhibiting the expression of nuclear factor κB (NF-κB).^13-15^ Measuring the levels of proinflammatory cytokines in COVID-19 patients, we have found a marked decrease in the levels of IL-6 in the LMWH treated patients compared to the patients without LMWH treatment (*p*<0·001), consistent with the proposed protective effect of LMWH. Changes in other inflammatory factors were not statistically significant. In addition, IL-6 can bind to HS on the cell surface, yielding a sufficiently high local concentration to activate signaling receptors,^40^ protect them against proteolysis, and promote paracrine action.^18,41^ An earlier study has reported that heparin binds IL-6, with affinity much higher than that of HS,^18^ thereby reducing its availability to its receptor complex. It, therefore, appears that LMWH reduces both the release of IL-6 and its biological activity.

Collectively, LMWH not only improves the coagulation dysfunction of COVID-19 patients,^42^ but exerts an anti-inflammatory effect by means of reducing IL-6 and increasing LYM%. We, therefore, favor the use of LMWH as a potential therapeutic drug for the treatment of COVID-19. We also suggest that non-anticoagulant species of LMWH that can be applied at high doses should be considered as a complement to conventional LMWH. To further support this conclusion, we are conducting a prospective clinical study to evaluate the efficacy and safety of one LMWH (enoxaparin) in the treatment of hospitalized adult patients with COVID-19 (Chinese Clinical Trial Registry, number:chiCTR2000030700), with the objective of providing a more powerful reference for the treatment conditions.

### Limitations

This study has several limitations. First, due to the retrospective design, we were unable to control the time intervals between examinations of the various indices in patients and the LMWH treatment schedule. Likewise, we could not estimate and manage the effective dose and timing of LMWH. Second, there were no critical cases in the two groups of patients; the treatment outcome of all cases was improvement and discharge, and there were no deaths. Finally, the findings are limited by the sample size and single-center design of our study.

## Contributors

CS, JPL and YZ conceptualized and designed the study, and CS and YZ had full access to all data, and took responsibility for data integrity and accuracy of the analysis. CS, CW, HX, JPL and IV wrote the manuscript. CY, FC and FZ reviewed the manuscript. FC, YH, TT and BD performed the statistical analysis. All authors contributed to data acquisition, analysis and interpretation, and approved the final version for submission.

## Data Availability

The data used to support the findings of this study are included within the article.

## Declaration of Interest

All authors declare no competing interests.

### Acknowledgements

This work was supported by the National Natural Science Foundation of China (No. 81603037 to SC) and the National Key Research and Development Plan of China(2017YFC0909900).

## Patient consent for publication

Not required

## Ethics approval

The human study was approved the Research Ethics Committee of Union Hospital, Tongji Medical College, Huazhong University of Science and Technology.

